# To ascertain the awareness among triage staff about clinical conditions treated in the Department of Hand Surgery from their referral pattern : A cross sectional observational study in a tertiary care center

**DOI:** 10.1101/2023.02.23.22281182

**Authors:** Maithreyi Sethu, Srinivasan Rajappa

**Affiliations:** Department of Hand Surgery, Sri Ramachandra Institute of Higher Education and Research, No 1, Ramachandra Nagar, Sri RamachandraNagar, Chennai, Tamil Nadu, India – 600116; Department of Hand Surgery, Sri Ramachandra Institute of Higher Education and Research, No 1, Ramachandra Nagar, Sri Ramachandra Nagar, Chennai, Tamil Nadu, India – 600116

**Keywords:** Triage, hand surgery department, hand surgery referral, triaging staff, out patient referral

## Abstract

**Objectives:**

- To ascertain the referral pattern and the awareness about clinical conditions treated in the Department of Hand Surgery among the triage staff.
- To determine if is there a difference between the triaging by the doctors and by the junior (less than 5 years in service) and the senior nurses (more than 5 years in service) with reference to clinical conditions which are treated in the Hand surgery Department.

**Materials and methods:** This was a cross-sectional observational study which was conducted in a tertiary care center in India. The study involved a total of 115 participants with junior nurses, senior nurses and doctors working in the triage service of the hospital.

All participants were shown a semi-structured tool with clinical scenarios which had photographs and clinical information in text, on a computer screen using Microsoft PowerPoint. The clinical scenario was also narrated to them. The referral pattern was noted down and tabulated.

**Results:** It is interesting to note that,

- Only in 1 out of the 20 scenarios, hand surgery was the most frequently referred department.
- Most of the Bony injuries of the hand and the forearm were mainly referred to orthopaedics.
- Burns and raw areas of the forearm were frequently referred to plastic surgery.
- Hand deformities were commonly referred to either plastic surgery or orthopaedics.
- Nerve injuries were referred to neurology and orthopaedics typically.
- Orthopaedics was the most referred department.
- The average reference percentage of the junior nurse was 3.91%, senior nurse was 21.9% and doctors was 23.95%. Junior nurses performed exceedingly poor in referring cases to the department of hand surgery as compared to the other two subgroups.

**Conclusion:** This study shows us the drawbacks in our present triaging system emphasizing the need for training our staff on new departments referral pattern, the advancements, and updated protocols.

## Background

Triage, derived from the French word trier meaning ‘to sort,’ is a process of prioritisation^1^. Though triage has its origins in military history, today it is used in a variety of medical settings^2^ from sorting/referring the patients at the reception or in the Emergency department to prioritising patients at the site of mass causality. Effective triage depends on careful choice as to who should make triage decisions^3^. It is optimal when the staff allocated for triaging are trained for it. A triage system must not only minimize risks to patient safety and misallocation of resources but must triage patients appropriately in an efficient manner^4^.

Recognition of the specialty of hand surgery and knowledge about the spectrum of cases it deals with seems to be in its infancy among the health professionals in India. Referrals to hand surgery department depend on the knowledge of staff handling triage in that institution. If the awareness of triage staff about Hand surgery and the spectrum of work which hand surgeons do is inadequate, this would result in inappropriate referrals of patients to other departments. This practice would in turn affect the growth and progress of the Hand surgery department.

## Method

This was a cross-sectional observational study which was conducted in a tertiary care center in India between January 2022 and February 2022. The study involved a total of 115 participants who consisted of junior nurses (experience less than 5 years), senior nurses (experience more than 5 years) and doctors working in the triage service of the hospital. The study was conducted in small batches of 3 to 7 volunteers. They were informed that it was a voluntary assessment study and were given the option of not participating. Consent was taken from the ones who had volunteered and at no point was it revealed to them that the study was conducted by the Department of Hand surgery to reduce the bias. Their personal information which would lead to their identification was not revealed in any part of the study.

All participants were shown a semi-structured tool with 25 clinical scenarios which had photographs and clinical information in text, on a computer screen using Microsoft PowerPoint. The clinical scenario was also narrated to them. Out of the 25 clinical scenarios, 20 pertained to clinical conditions which were treated in the department of Hand surgery. Consensus on which is the ideal department to send the patients to in these 25 clinical scenarios in this institution was determined by a panel of intra-institutional experts, which involved three plastic surgeons and two orthopedicians from the institution.

The participants were shown each clinical scenario at one minute intervals. During this minute each volunteer had to respond by writing down which department they would refer the patient to first.

The responses were tabulated in an excel sheet and statistical analysis was used to calculate the mean, standard deviations and percentages.

1. For each of the clinical scenario, the three most frequently marked departments to which the triaging staff would refer the patients to was calculated as percentage.
2. The participants were then divided into junior nurses, senior nurses and doctors. And the correct responses (correctly referring the patient to the department of hand surgery) given by each group was calculated as a percentage.
3. The difference in this percentage of responses (referring the patient to the department of hand surgery) between the doctors, junior and senior nurses were compared.

### Inclusion Criteria

- All the nurses and doctors (which included House surgeons/CRRI and first year post graduates from different specialities) in the triaging service area of the tertiary care hospital

### Exclusion

- Junior nurses with less than 1 month of experience at the triage
- Non-medical staff from the administrative wing
- Staff who were aware about this triage study
- Staff in the emergency department
- Staff in the reception area of the different subspecialities

The Institutional Ethics Committee approval was sought before starting the study, IEC-NI/21/OCT/80/136.

## Results

The study involved 115 participants (triaging staff) with 36 doctors and 79 nurses. Out of the 79, 38 were junior nurses and 41 were senior nurses. Most of the doctors were fresh graduates within 2 years of completing their under-graduation. Most of the nurses were females. The sex distribution was better with doctors. The average age of nurses was 28.9 years while that of the doctors was 23.4 years. The demography of the volunteers has been tabulated in Table 1.

**Table 1.** Demography of the volunteers

| Variable | Nurses (N=79) | Doctors (N=36) |
| --- | --- | --- |
| Age (years) | 28.9+/- 6.9 | 23.4 +/- 1.4 |
| Gender |  |  |
| Male | 1 (1.2%) | 20 (55.5%) |
| Female | 78 (98.7%) | 16 (44.4%) |
| Experience |  |  |
| <5yrs | 41 | 36 |
| >5yrs | 38 | 0 |
| In months | 80.2 +/- 4.4 | 6.0 +/- 1.7 |

The three most frequently marked departments to which the triaging staff would refer the patients to was calculated as percentages (Table 2). To summarize the results, it was found that in none of the 20 clinical conditions Hand Surgery made it as the leading choice of referral except in a clinical scenario of ulnar nerve injury. But even in this case it shared the first place with orthopaedics.

**Table 2.** Top three responses to each of the 20 clinical conditions

| Diagnosis & Department to be referred | Rank | Most referred Departments | Numbers | Percentage |
| --- | --- | --- | --- | --- |
| 1. Proximal phalanx Fracture | 1 | Orthopedics | 78 | 64.40% |
| Hand Surgery | 2 | Hand Surgery | 27 | 22.30% |
|  | 3 | Neurology | 4 | 3.30% |
| 2. Stroke | 1 | Neurology | 81 | 66.90% |
| Neurology | 2 | Medicine | 23 | 19.00% |
|  | 3 | Orthopedics | 4 | 3.30% |
| 3. Hand-Burns | 1 | Plastic Surgery | 43 | 35.10% |
| Hand Surgery | 2 | Paediatrics | 36 | 29.70% |
|  | 3 | ER | 11 | 9.00% |
| 4. Elbow Dislocation | 1 | Orthopedics | 100 | 82.60% |
| Orthopedics | 2 | Hand Surgery | 6 | 4.90% |
|  | 3 | Sports Medicine | 5 | 4.10% |
| 5. Flexor Tenosynovitis | 1 | Medicine | 32 | 26.40% |
| Hand Surgery | 2 | Surgery | 28 | 23.10% |
|  | 3 | Hand Surgery | 24 | 19.80% |
| 6. Flexor Tendon Injury | 1 | Orthopedics | 53 | 43.80% |
| Hand Surgery | 2 | Hand Surgery | 38 | 31.40% |
|  | 3 | Physiotherapy | 9 | 7.40% |
| 7. Scaphoid Fracture | 1 | Orthopedics | 75 | 61.90% |
| Hand Surgery | 2 | Hand surgery | 21 | 17.30% |
|  | 3 | Physiotherapy | 6 | 4.90% |
| 8. Carpel Tunnel Syndrome | 1 | Medicine | 38 | 31.40% |
| Hand Surgery | 2 | Neurology | 37 | 30.50% |
|  | 3 | Hand Surgery | 12 | 9.90% |
| 9. Rheumatic Hand Deformity | 1 | Hand Surgery | 27 | 22.30% |
| Rheumatology | 2 | Rheumatology | 27 | 22.30% |
|  | 3 | Orthopedics | 26 | 21.40% |
| 10. Ulnar Nerve Injury | 1 | Hand Surgery | 35 | 28.90% |
| Hand Surgery | 2 | Orthopedics | 35 | 28.90% |
|  | 3 | Neurology | 17 | 14.00% |
| 11. Obstetric Brachial Plexus Palsy | 1 | Orthopedics | 43 | 35.50% |
| Hand Surgery | 2 | Paediatrics | 40 | 33.00% |
|  | 3 | Neurology | 16 | 13.20% |
| 12. (Cleft) Hand Deformity | 1 | Orthopedics | 55 | 45.40% |
| Hand surgery | 2 | Plastic Surgery | 29 | 23.90% |
|  | 3 | Hand Surgery | 11 | 9% |
| 13. Traumatic (Metacarpal) Hand Deformity | 1 | Plastic Surgery | 62 | 51.20% |
| Hand Surgery | 2 | Hand Surgery | 34 | 28.00% |
|  | 3 | Orthopedics | 12 | 9.90% |
| 14. Venflon site Ulcer – Dorsum of hand | 1 | Surgery | 63 | 52.00% |
| Hand Surgery | 2 | Medicine | 27 | 22.30% |
|  | 3 | Hand Surgery | 8 | 6.60% |
| 15. Vascular Malformation of hand | 1 | Vascular Surgery | 60 | 49.50% |
| Hand Surgery | 2 | Hand Surgery | 18 | 14.80% |
|  | 3 | Surgery | 13 | 10.70% |
| 16. Cerebral Palsy | 1 | Neurology | 32 | 26.40% |
| Neurology | 2 | Orthopedics | 26 | 21.40% |
|  | 3 | Paediatrics | 20 | 16.50% |
| 17. Elbow Sprain | 1 | Orthopedics | 100 | 82.60% |
| Hand Surgery | 2 | Hand Surgery | 4 | 3.30% |
|  | 3 | Physiotherapy | 2 | 1.60% |
| 18. Carcinoma (Squamous cell) little finger | 1 | Oncology | 53 | 43.80% |
| Hand Surgery | 2 | Hand Surgery | 21 | 17.30% |
|  | 3 | Surgery | 20 | 16.50% |
| 19. Shoulder Periarthritis | 1 | Orthopedics | 70 | 57.80% |
| Orthopedics | 2 | Medicine | 23 | 19.00% |
|  | 3 | Physiotherapy | 5 | 4.10% |
| 20. Thoracic Outlet Syndrome | 1 | Neurology | 39 | 32.20% |
| Hand Surgery | 2 | Medicine | 26 | 21.40% |
|  | 3 | Orthopedics | 22 | 18.10% |
| 21. Volkmann Ischemic Contracture | 1 | Orthopedics | 48 | 39.60% |
| Hand Surgery | 2 | Hand Surgery | 29 | 23.90% |
|  | 3 | Neurology | 15 | 12.30% |
| 22. Raw Area Forearm (skin loss) | 1 | Plastic Surgery | 41 | 33.80% |
| Hand Surgery | 2 | Surgery | 30 | 24.70% |
|  | 3 | Orthopedics | 19 | 15.70% |
| 23. Colles' Fracture | 1 | Orthopedics | 90 | 74.30% |
| Hand Surgery | 2 | Hand Surgery | 18 | 14.80% |
|  | 3 | Medicine | 2 | 1.60% |
| 24. Traumatic Brachial Plexus Palsy | 1 | Neurology | 51 | 42.10% |
| Hand Surgery | 2 | Orthopedics | 45 | 37.10% |
|  | 3 | Hand Surgery | 8 | 6.60% |
| 25. Congenital (Madelung) Hand deformity | 1 | Orthopedics | 62 | 51.20% |
| Hand Surgery | 2 | Hand Surgery | 33 | 27.20% |
|  | 3 | Neurology | 8 | 6.60% |

- Questions 2,4,9, 16 and 19 marked in Red are Non-hand surgery referral cases added to questionnaire

It is interesting to note that,

- Most of the Bony injuries of the hand and the forearm were mainly referred to Orthopaedics.
- Burns and raw areas of the forearm were referred to plastic surgery.
- Problems with soft-tissue injury such as flexor tendon injury and Volkmann’s Ischemic contracture were also referred to orthopaedics.
- When terms like small vascular lesion or tumours/growth were used, patients were often referred to vascular surgery and onco-surgery department, without much attention being given to the entire clinical scenario.
- Hand deformities were referred to either plastic surgery (Cleft hand deformity) or orthopaedics (Madelung deformity).
- Nerve injuries such as Thoracic outlet syndrome and traumatic brachial plexus palsy were referred to Neurology, whereas cut injury of nerve in the forearm and obstetric brachial plexus palsy were referred to orthopaedics.
- When the patient had swelling or weakness of the forearm such as in flexor tenosynovitis or carpal tunnel syndrome but the history showed associated diabetes, fever or hypothyroidism, most of the patients were referred to the department of medicine.
- Orthopaedics was the most referred department.
- Physiotherapy also made it to the top three for conditions such as flexor tendon injury, scaphoid fracture, elbow pain and periarthritis shoulder (6,7,17 & 19).

For closer analysis, the responses were divided as given by the different sub-groups-doctors, junior nurses and senior nurses. The number and percentage of the responses appropriately referring the patient to the department of hand surgery by each of these subgroups were calculated (Table 3).

**Table 3.**
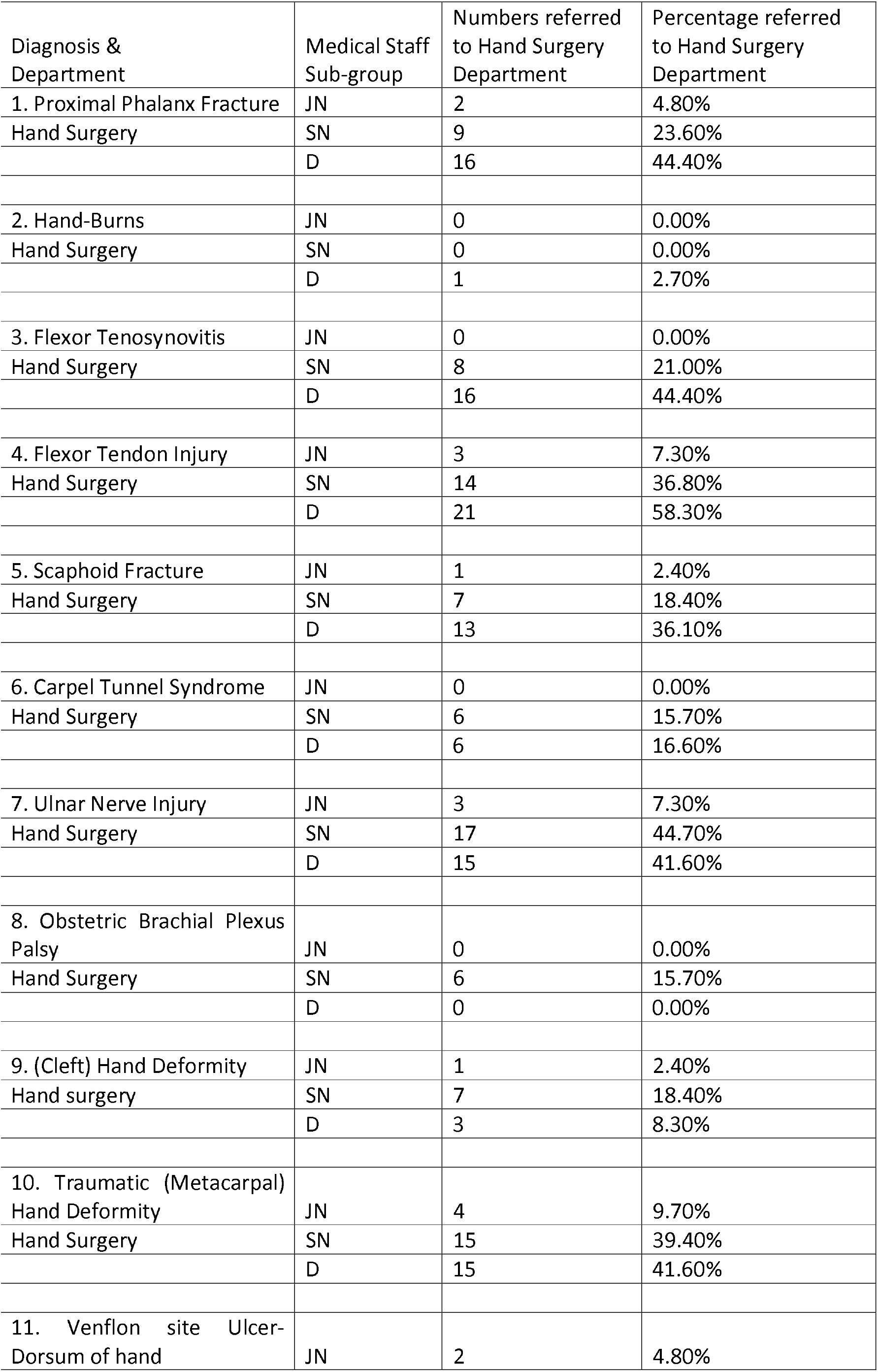
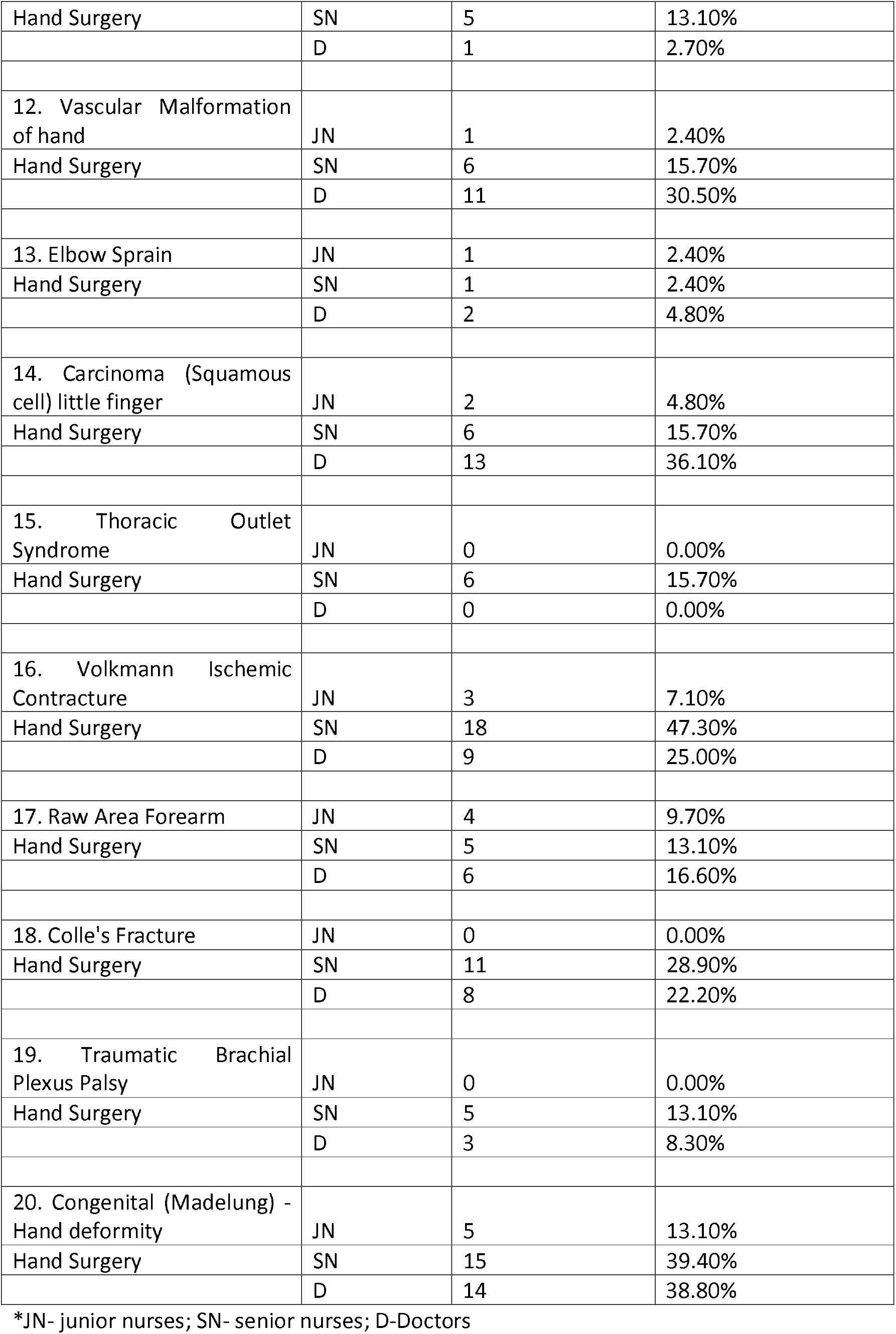
Comparison of responses of the 20 clinical conditions between doctors, junior nurses and senior nurses

| Diagnosis & Department | Medical Staff Sub-group | Numbers referred to Hand Surgery Department | Percentage referred to Hand Surgery Department |
| --- | --- | --- | --- |
| 1. Proximal Phalanx Fracture | JN | 2 | 4.80% |
| Hand Surgery | SN | 9 | 23.60% |
|  | D | 16 | 44.40% |
| 2. Hand-Burns | JN | 0 | 0.00% |
| Hand Surgery | SN | 0 | 0.00% |
|  | D | 1 | 2.70% |
| 3. Flexor Tenosynovitis | JN | 0 | 0.00% |
| Hand Surgery | SN | 8 | 21.00% |
|  | D | 16 | 44.40% |
| 4. Flexor Tendon Injury | JN | 3 | 7.30% |
| Hand Surgery | SN | 14 | 36.80% |
|  | D | 21 | 58.30% |
| 5. Scaphoid Fracture | JN | 1 | 2.40% |
| Hand Surgery | SN | 7 | 18.40% |
|  | D | 13 | 36.10% |
| 6. Carpel Tunnel Syndrome | JN | 0 | 0.00% |
| Hand Surgery | SN | 6 | 15.70% |
|  | D | 6 | 16.60% |
| 7. Ulnar Nerve Injury | JN | 3 | 7.30% |
| Hand Surgery | SN | 17 | 44.70% |
|  | D | 15 | 41.60% |
| 8. Obstetric Brachial Plexus Palsy | JN | 0 | 0.00% |
| Hand Surgery | SN | 6 | 15.70% |
|  | D | 0 | 0.00% |
| 9. (Cleft) Hand Deformity | JN | 1 | 2.40% |
| Hand surgery | SN | 7 | 18.40% |
|  | D | 3 | 8.30% |
| 10. Traumatic (Metacarpal) Hand Deformity | JN | 4 | 9.70% |
| Hand Surgery | SN | 15 | 39.40% |
|  | D | 15 | 41.60% |
| 11. Venflon site Ulcer-Dorsum of hand | JN | 2 | 4.80% |
| Hand Surgery | SN | 5 | 13.10% |
|  | D | 1 | 2.70% |
| 12. Vascular Malformation of hand | JN | 1 | 2.40% |
| Hand Surgery | SN | 6 | 15.70% |
|  | D | 11 | 30.50% |
| 13. Elbow Sprain | JN | 1 | 2.40% |
| Hand Surgery | SN | 1 | 2.40% |
|  | D | 2 | 4.80% |
| 14. Carcinoma (Squamous cell) little finger | JN | 2 | 4.80% |
| Hand Surgery | SN | 6 | 15.70% |
|  | D | 13 | 36.10% |
| 15. Thoracic Outlet Syndrome | JN | 0 | 0.00% |
| Hand Surgery | SN | 6 | 15.70% |
|  | D | 0 | 0.00% |
| 16. Volkmann Ischemic Contracture | JN | 3 | 7.10% |
| Hand Surgery | SN | 18 | 47.30% |
|  | D | 9 | 25.00% |
| 17. Raw Area Forearm | JN | 4 | 9.70% |
| Hand Surgery | SN | 5 | 13.10% |
|  | D | 6 | 16.60% |
| 18. Colle's Fracture | JN | 0 | 0.00% |
| Hand Surgery | SN | 11 | 28.90% |
|  | D | 8 | 22.20% |
| 19. Traumatic Brachial Plexus Palsy | JN | 0 | 0.00% |
| Hand Surgery | SN | 5 | 13.10% |
|  | D | 3 | 8.30% |
| 20. Congenital (Madelung) - Hand deformity | JN | 5 | 13.10% |
| Hand Surgery | SN | 15 | 39.40% |
|  | D | 14 | 38.80% |

\*JN- junior nurses; SN- senior nurses; D-Doctors

In all the 20 questions, except for doctors in the clinical scenario of flexor tendon injury (58%) none of the 3 subgroups referred even half (50%) of the patients to Hand surgery.

There were several questions such as scalding burns of hand, flexor tenosynovitis, carpal tunnel syndrome, obstetric brachial plexus palsy, thoracic outlet syndrome, Colles fracture and traumatic brachial plexus palsy where one or more sub-groups did not refer any patients to the department of hand surgery.

The average reference percentage of the junior nurse was 3.91%, senior nurse was 21.9% and doctors was 23.95%. Junior nurses performed exceedingly poor in referring cases to the department of hand surgery as compared to the other two subgroups.

## Discussion

Hand surgery is as old as surgery itself, dating back to prehistoric times, or to Egyptian and Greek antiquity. Hippocrates (460±356 BC) has given useful advice about reduction and stabilization of wrist, hand and finger fractures and dislocations^5^.

Even though Hand surgery had existed for such a long time, it required an eye opener like Dr. Sterling Bunnell, the American father of Hand Surgery to establish a separate department for it. He trained several hand surgeons and helped organise several hand surgery centers following the period of the second world war^6 7^ which played a major role in giving hand surgery an opportunity to crown herself as a unique and separate speciality. Ironically even after several decades of existence, knowledge and understanding of common hand surgical conditions and awareness of the subspeciality among physicians in India is still negligible^8^. The poor literacy level and level of awareness of the general public in India is also a setback. With this in one side, on the other extreme there are views from leading scholars that the expanding knowledge base of Hand surgery, increasing technological advances and changes in the surgical experiences makes a 12-month experience too short even for a hand surgery trainee^9^.

Doctors from other specialities are themselves quite unaware as to the spectrum of patients catered by the department of hand surgery. It is no surprise that junior residents and nursing staff are also in the same milieu. This study clearly shows the lack of awareness and knowledge about the department among the triaging staff who are made up of junior doctors and nurses.

The lack of awareness in the triaging staff could be the starting point of a vicious cycle (lack of awareness ⟶ failure to refer patients ⟶ decrease in patient load ⟶ department not achieving its full potential⟶ lack of awareness in other hospital staff) which could interfere with the establishment of the speciality. Since proper triaging regarding patients needing hand surgery becomes important for the patient to get the most appropriate treatment with minimum shunting around but also helps in the improvement and refinement of the speciality. Even in complicated, complex cases or patient operated unsuccessfully elsewhere, it is possible to enhance the skill and knowledge only if the patient reaches the department promptly. This is where triaging plays a major role and it becomes our responsibility to make the triaging staff, the paramedics and even the public acquainted with the conditions treated in the department of hand surgery.

In the fast-paced environment of the out patients’ department, critical thinking, evaluation and decision making by the triaging staff plays a key role in efficient treatment of patients^10^. It requires the staff to have a strong foundation in basic clinical knowledge which needs to be frequently updated and refreshed equipping them to make quick logical decisions. Not many institutions have a comprehensive standardised training programme for triaging, thus creating a lacuna in the process. This study further emphasises the above-mentioned fact.

For triage training, a combination of multiple teaching methods and training approaches such as human patient simulation, computer learning games or virtual reality triage training might offer an alternative or adjunct for triage training in non-clinical situations^11^. However, these would be much more labor-intensive than currently used methods which include regular refresher training programs, testing of factual knowledge (e.g. with case scenarios), and direct observation of triage performance including feedback^11^.

## Conclusion

Triage systems aim not only to ensure clinical justice for the patient, but also to provide an effective tool for departmental organisation, monitoring and evaluation^12^.

From the study it is clear that most of the triaging staff were unaware of the spectrum of patients being dealt with in the department of hand surgery. They were mostly referring the patients to department to which they would have referred the patients to if the dept of hand surgery did not exist. Junior nurses performed sub-standard to the senior nurses and doctors who were also struggling to identify the department of reference for the patients.

This study shows us the drawbacks in our present triaging system causing some of the problems a new budding department would face in its blooming stages such as low patient load, increased number of references of a patient, decreased cure rates and slow growth of the department. We focus on propagating the speciality or the medical condition via social media and posters to reach the public. But we have forgotten an important intermediate building block, the triaging staff who play a very important role of delegating the patient to the correct department. Along with creating public awareness, training our staff on the advancements and updated protocols is a very crucial step in the development of an institution and a healthy society.

### Limitation of the study

This is only the first part of a larger study. A training module is to be conducted for the triaging staff following which a post training assessment will be done to see the efficiency of the training and if required modification of the training programme will be done. Further studies can also be done to compare different triage algorithms to help overcome the variability and subjectivity.

## Supporting information

proforma

## Data Availability

All data produced in the present study are available upon reasonable request to the authors

## Acknowledgements

None

