## Supplementary material for "To ascertain the awareness among triage staff about clinical conditions treated in the Department of Hand Surgery from their referral pattern : A cross sectional observational study in a tertiary care center": proforma

STAFF NAME

ID NUMBER –

AGE –

SEX –

Contact number-

Educational qualification-

Experience in triage team-

Experience after MBBS-

| Question Number | Department Referred to | Confidence  (1-10) |
| --- | --- | --- |
| 1 |  |  |
| 2 |  |  |
| 3 |  |  |
| 4 |  |  |
| 5 |  |  |
| 6 |  |  |
| 7 |  |  |
| 8 |  |  |
| 9 |  |  |
| 10 |  |  |
| 11 |  |  |
| 12 |  |  |
| 13 |  |  |
| 14 |  |  |
| 15 |  |  |
| 16 |  |  |
| 17 |  |  |
| 18 |  |  |
| 19 |  |  |
| 20 |  |  |
| 21 |  |  |
| 22 |  |  |
| 23 |  |  |
| 24 |  |  |
| 25 |  |  |

I AM WILLING TO TAKE PART IN THE STUDY

SIGNATURE
